# Risk of progression to pulmonary tuberculosis among household contacts with chest radiographic abnormalities in South Africa

**DOI:** 10.64898/2026.06.01.26354586

**Authors:** Humphrey Mulenga, Evans Muchiri, Simon C. Mendelsohn, Stephanus T. Malherbe, Tumelo Moloantoa, Michele Tameris, Fernanda Maruri, Firdows Noor, Ravindre Panchia, Khuthadzo Hlongwane, Kim Stanley, Kate Hadley, Neil Martinson, Gerhard Walzl, Thomas J. Scriba, Timothy R. Sterling, Mark Hatherill, the RePORT South Africa Study Team

## Abstract

**Background:** High-risk subgroups among household contacts of persons with tuberculosis (TB) might benefit from additional interventions. However, the significance of an abnormal baseline chest radiograph (CXR) suggestive of TB, despite negative sputum microbiology, is uncertain.

**Methods:** Adults (≥18 years) with recent household TB exposure were enrolled at three South African sites (April 2021– September 2022). All participants underwent symptom screening, CXR, and sputum Xpert Ultra and MGIT culture. Pulmonary TB diagnosis was microbiologically-confirmed. Participants were followed for symptomatic incident TB through 12 months. Multivariable logistic regression identified factors associated with abnormal CXR suggestive of TB. Poisson regression estimated incidence rate ratios (IRR).

**Results:** Baseline CXR were abnormal in 157/795 (19.7%) participants; associated with older age (adjusted odds ratio, aOR=1.04, 95%CI 1.02–1.05); prior TB (aOR=6.39, 95%CI 4.18–9.78); and current smoking (aOR=1.61, 95%CI 1.00–2.62). Symptomatic incident TB developed in 8/795 (1.0%) participants, including 7/8 (87.5%) who were asymptomatic and 4/8 (50.0%) with abnormal CXR at baseline. TB incidence was four-fold higher in those with abnormal versus normal CXR (IRR=4.02, 95%CI 1.01–15.97), with a risk difference of 1,969 (95%CI -657– 4,595) per 100,000 person-years, but after median 12.1 (IQR 11.1–13.1) months follow-up, 153/157 (97.5%) had not progressed to incident TB.

**Conclusions:** Adult contacts with CXR abnormalities, but without prevalent TB, had a four-fold higher risk of TB within one year, compared to those with normal CXR. This additional risk warrants targeted preventive treatment and extended surveillance, but since most remained TB-free, therapeutic TB treatment is not justified.

**Key points:** Household contacts with abnormal CXR suggestive of TB have four-fold increased risk of incident TB disease, compared to normal CXR, which justifies targeted preventive treatment and extended surveillance, but since the majority remain healthy, therapeutic TB treatment is not warranted.

## Introduction

Tuberculosis (TB) remains a major global health challenge. In 2024, an estimated 10.7 million people developed TB and approximately 1.25 million died [1]. Achieving End TB Strategy targets of a 95% reduction in TB deaths and a 90% reduction in TB incidence rate by 2035 will require prompt diagnosis and effective treatment of TB disease, alongside targeted prevention for people at highest risk of disease progression [2].

Household contacts of patients with pulmonary TB (PTB) are at elevated risk of *Mycobacterium tuberculosis* (Mtb) infection and TB disease, particularly in the first 1–2 years after exposure [3, 4]; therefore, systematic screening is recommended by TB control programmes [5]. Screening commonly includes symptom assessment and chest radiography (CXR), which, if positive, trigger sputum microbiological testing to confirm and treat TB disease. However, this diagnostic algorithm would exclude asymptomatic individuals with CXR abnormalities suggestive of TB who have negative initial sputum Mtb culture or nucleic acid amplification tests, despite meeting international consensus on early TB (ICE-TB) criteria for asymptomatic (previously termed subclinical), non-infectious TB [6]. These individuals may be at greater risk of progressing to symptomatic, infectious disease [7].

Functional imaging modalities such as FDG-PET/CT can identify metabolically active lesions and have been shown to detect TB disease earlier, when conventional microbiological tests are negative, highlighting limitations of current diagnostics for early or paucibacillary TB [8]. A Cochrane systematic review and meta-analysis showed that approximately 11% of the general population screened in high-burden settings have CXR abnormalities suggestive of TB without microbiological confirmation [9]; and a mass TB screening study in China reported 16-fold increased risk of incident TB in such individuals with CXR suggestive of active TB [10]. Further, in a Peruvian study, half of all household contacts 15 years and older who developed incident TB had CXR abnormalities at baseline; and among those who were asymptomatic, abnormal CXR was associated with 15-fold increased risk of progression to TB [11]. These estimates are consistent with annualized risk of progression from microbiologically negative to positive TB disease, in participants with baseline CXR suggestive of inactive or active TB, ranging from 1-10%, respectively [7]. Despite this apparent elevated risk of TB [7], clinical management guidelines for individuals with CXR abnormalities suggestive of TB, particularly those without symptoms, are not well defined.

While TB preventive treatment (TPT) substantially reduces disease progression in high-risk groups, including household contacts and people living with HIV (PLWH) [12], the very high background prevalence of interferon-γ release assay (IGRA) positivity—up to 80% in some South African communities [13]—limits the utility of IGRA as a tool to guide therapeutic decision-making. Accordingly, TPT is recommended for all household contacts of known TB patients, regardless of IGRA status. However, the magnitude of additional risk associated with abnormal baseline CXR findings, when baseline symptoms are absent and sputum microbiology is negative, is unclear, but this risk may warrant additional interventions. Better characterisation of this group is needed to inform risk-benefit stratification for evidence-based, therapeutic decision-making.

We evaluated the prevalence of baseline CXR abnormalities suggestive of TB; associated risk factors such as symptom and IGRA status; 12-month incidence of microbiologically-confirmed TB; and the risk of incident TB within one year among adult household contacts of a known TB patient in a South African cohort. We hypothesised that abnormal baseline CXR findings would identify a subgroup of household contacts at substantially higher risk of incident TB that might warrant a therapeutic, rather than preventive, treatment regimen.

## Methodology

### Study design and participants

The Regional Prospective Observational Research for Tuberculosis (RePORT) South Africa network enrolled participants in a prospective observational cohort study, as previously described [14.]. Briefly, adults (≥18 years) with household exposure within the past six months to an adult index patient with untreated or inadequately treated PTB were enrolled at three centres in South Africa (Worcester and Ravensmead, Western Cape Province; Klerksdorp, Northwest Province). Exposure was defined as sleeping in the same household or four or more hours of other close exposure per week. Participants were excluded if they were unlikely to attend study visits or had any condition that might interfere with their ability to provide informed consent or adhere to study requirements, including alcohol or drug dependence and incarceration.

All participants were systematically investigated for prevalent TB disease at baseline, regardless of symptom or CXR screening results. Baseline TB investigations included symptom screening, CXR, IGRA (QuantiFERON-TB Gold Plus, QIAGEN, Germany), and collection of a single spontaneously expectorated sputum sample for Xpert Ultra (Cepheid, USA) and liquid culture (Mycobacteria Growth Indicator Tube [MGIT], BACTEC, Beckton Dickinson, USA). Participants unable to produce sputum were classified as microbiologically negative. Induced sputum was not performed.

The microbiological reference standard (MRS) for confirmed TB disease was at least one sputum specimen positive by MGIT culture or Xpert Ultra (excluding trace-positive results). Prevalent TB was diagnosed on sputum samples collected within 30 days of baseline. A participant was classified as symptomatic if they reported one or more of cough, fever, weight loss, fatigue, night sweats, pleuritic chest pain, or haemoptysis, of any duration. CXR were read once by an investigator at each site using a standardised form; abnormal CXR were defined by the presence of any radiographic features suggestive of TB involving the lungs, pleura, or mediastinum, including infiltrates, opacities, cavities, nodules, fibrosis, pleural effusion, or lymphadenopathy. IGRA testing was performed according to the manufacturer’s instructions, with an interferon-γ response ≥0.35 IU/mL considered positive.

Participants without prevalent TB were followed for 12 months for microbiologically-confirmed incident TB, defined as TB disease diagnosed on sputum collected after 30 days from baseline. Follow-up TB investigations were symptom-triggered; participants reporting symptoms at the month 6 or month 12 visits provided sputum for Xpert Ultra and MGIT culture. All individuals diagnosed with microbiologically-confirmed TB, or with a clinical diagnosis of TB based on symptoms or CXR abnormalities suggestive of active disease, were referred for therapeutic treatment.

### Statistical analysis

Statistical analyses were conducted in Stata (version 17, Stata Corp LLC, USA). All statistical tests were two-sided and p values <0.05 were considered significant. Bivariate analyses of participant characteristics by CXR status used univariable logistic regression, with point estimates reported as odds ratio (OR) with 95% confidence interval (95%CI). Independent sociodemographic and clinical predictors of an abnormal baseline CXR were evaluated using multivariable logistic regression. The final model included all clinically important variables, plus those associated with abnormal CXR in univariable analyses, with results reported as adjusted odds ratio (aOR) and 95%CI.

To compare progression to incident TB among participants with abnormal versus normal baseline CXR, we estimated the incidence rate ratio (IRR), and absolute risk difference with 95%CI, using Poisson regression with log person-time as an offset and robust standard errors. Poisson regression was used to compute incident TB rates, given the low event frequency. The regression model was not adjusted for any covariate owing to the low absolute incident TB event frequency. Follow-up time accrued from the baseline visit to the earliest of: (i) incident TB (event), (ii) last study contact for participants lost to follow-up, (iii) death or withdrawal, or (iv) administrative censoring at 12 months. Participants with baseline microbiologically-confirmed TB, or unconfirmed TB treated on clinical grounds, or missing microbiology results, or having less than 31 days of follow-up time, were excluded from incident TB analyses.

The primary analysis used a strict definition of sputum microbiological negativity and only included participants with negative baseline sputum MGIT culture and Xpert Ultra results and an available CXR report. A sensitivity analysis included participants with a negative Xpert Ultra but no MGIT culture result, and participants with trace-positive Xpert Ultra and a negative MGIT culture result, provided a CXR was available. A second sensitivity analysis broadened the TB disease endpoint by including unconfirmed incident TB treated on clinical grounds.

### Ethical approvals

The study protocol was approved by the institutional research ethics committees at each participating South African site and Vanderbilt University Medical Center, USA. All participants provided written informed consent prior to participation.

## Results

In total, 979 household contacts were enrolled between April 2021 and September 2022 [14]. The primary analysis excluded 184 participants with prevalent TB (n=51) or uncertain TB status (n=82), no available CXR (n=39), or less than 30 days of follow-up from baseline (n=12). The primary analytic cohort comprised 795 participants with negative baseline sputum Xpert Ultra and MGIT culture results and an available CXR report (**Figure 1**).

**Figure 1.**
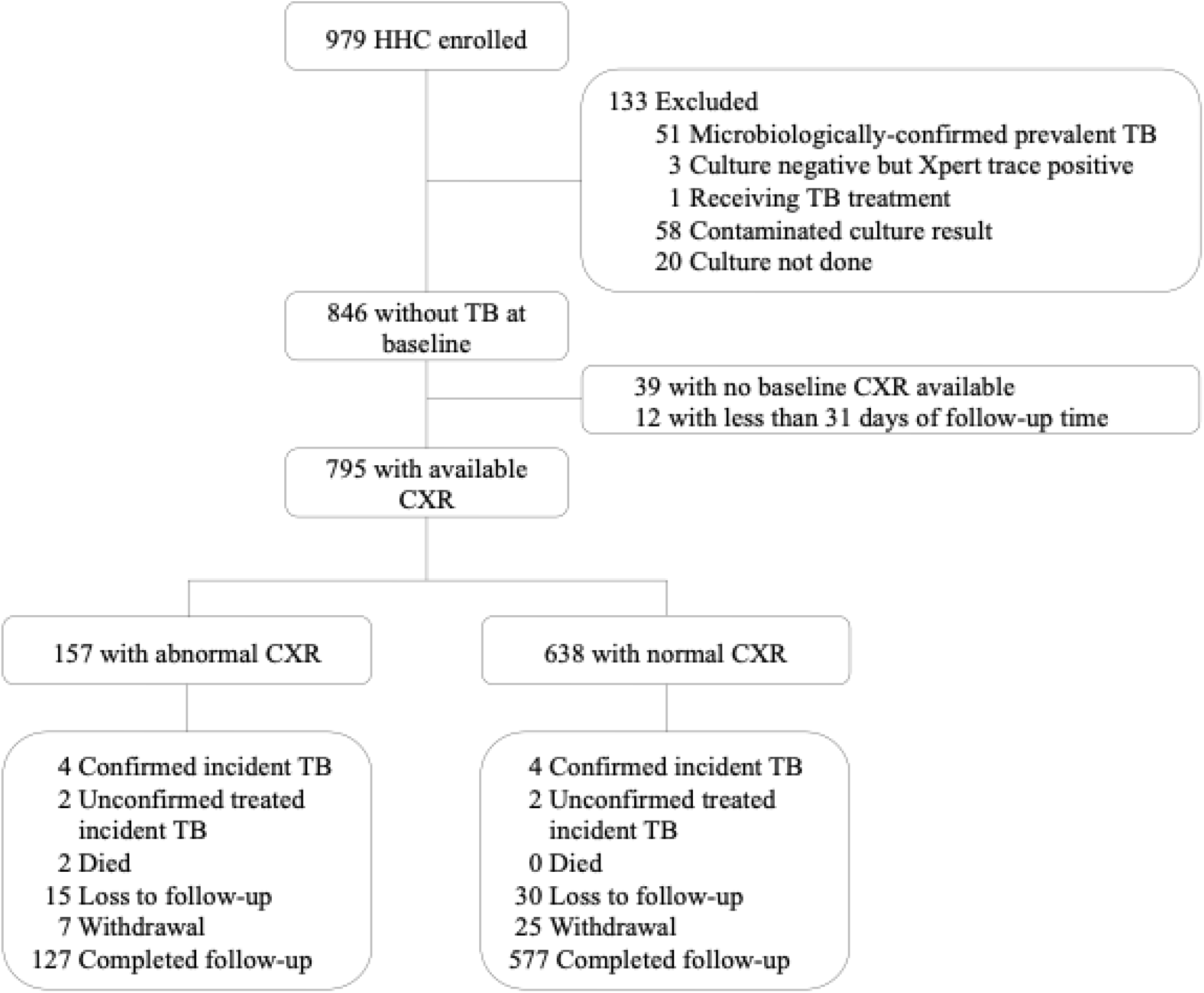
Study flow diagram. The study flow diagram shows exclusions, chest radiograph results, and follow-up outcomes for the primary analysis. Unconfirmed, treated incident TB denotes participants who were clinically diagnosed with TB without microbiological confirmation and who started therapeutic TB treatment. HHC, household contact. TB, tuberculosis. CXR, chest radiograph.

Median age was 34.8 years (IQR 25.4–48.0) and 508 (63.9%) of 795 participants were female. Prior TB was reported by 144 (18.1%); 150 (18.9%) were PLWH; 8 (1.0%) reported receiving TPT; and 677 (85.2%) were asymptomatic. Among 741 (93.2%) participants with a valid IGRA result, 579 (78.1%) were IGRA-positive (**Table 1**).

**Table 1.**
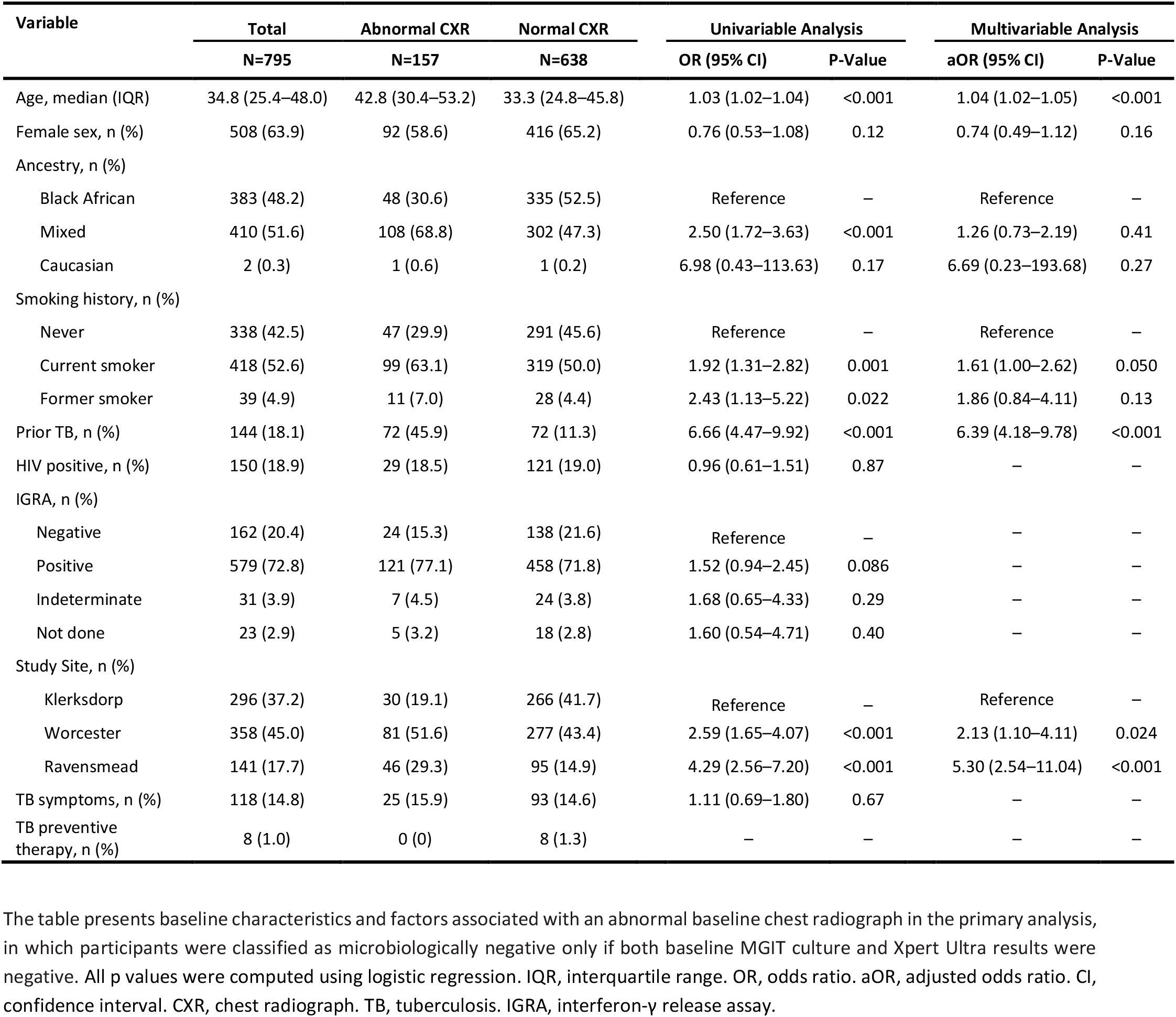
Baseline characteristics of the study population and factors associated with an abnormal baseline chest radiograph.

An abnormal CXR suggestive of TB was observed in 157 (19.7%) of 795 participants. In univariable analyses, participants with abnormal versus normal CXR were older (median 42.8 years vs 33.3 years; p<0.001); more likely to be of mixed ancestry (68.8% vs 47.3%; p<0.001); current (63.1% vs 50.0%; p=0.001) or former (7.0% vs 4.4%; p=0.022) smokers; and more likely to report prior TB (45.9% vs 11.3%; p<0.001). Baseline IGRA and TB symptom status did not differ between participants with abnormal and normal CXR (**Table 1**).

In multivariable logistic regression (after adjusting for study site, sex, and ancestry), increasing age (aOR 1.04 per year, 95%CI 1.02–1.05) and prior TB (aOR 6.39, 95%CI 4.18–9.78) were significantly associated with abnormal baseline CXR. Current smoking was weakly associated with abnormal baseline CXR (aOR 1.61, 95%CI 1.00–2.62).

In total, over median 11.9 (IQR 11.8–12.1) months, 45 participants were lost to follow-up; 32 withdrew; 2 died; 4 were treated for incident TB without microbiological confirmation; and 704 completed 12 months of follow-up without developing TB (**Figure 1**). Overall, 8/795 (1.0%) participants progressed to microbiologically-confirmed incident TB, corresponding to an incidence rate of 1,044 per 100,000 person-years (95%CI 322–1,766). Seven of the eight (87.5%) TB progressors had been asymptomatic at baseline (**Supplementary Table 1**).

Among the 157 participants with abnormal baseline CXR, median follow-up time was 12.1 (IQR 11.1–13.1) months. Four of 157 (2.5%) progressed to incident TB and 153 (97.5%) did not progress by the time of censoring or completion of follow-up. Among the 638 participants with normal baseline CXR, median follow-up time was 11.3 (IQR 10.9–12.5) months. Four of 638 (0.6%) progressed to incident TB and 634 (99.4%) did not progress by the time of censoring or completion of follow-up. Incident TB was more common among participants with abnormal CXR, compared to those with normal CXR (IRR 4.02, 95%CI 1.01–15.97), with incidence rates of 2,621 (95%CI 74–5,167) and 652 (95%CI 13–1,290) per 100,000 person-years, respectively. The risk difference for an abnormal CXR compared to a normal CXR was 1,969 (95%CI -657–4594; **Table 2**) per 100,000 person-years.

**Table 2.**
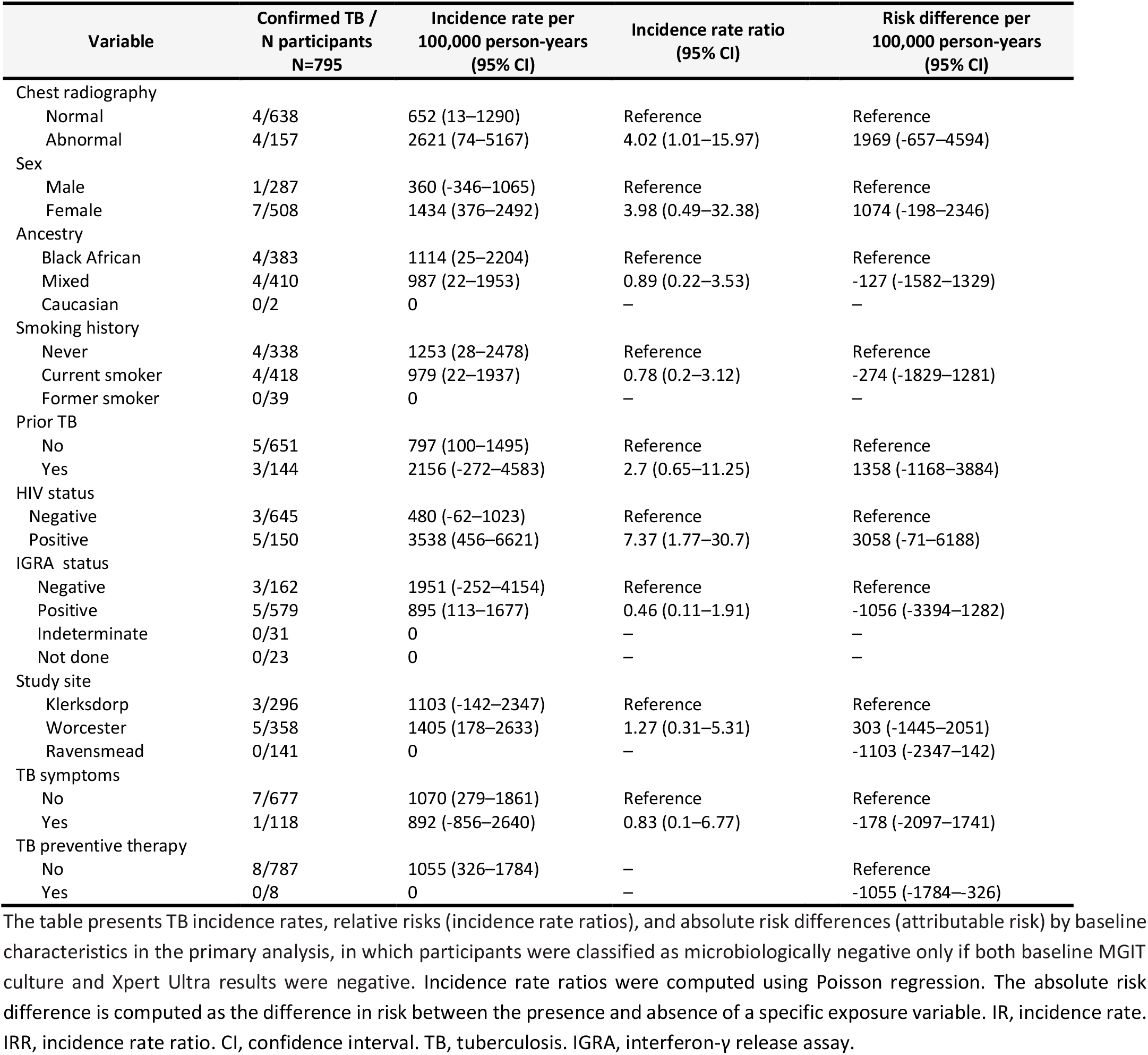
Risk of progression to incident pulmonary TB by baseline characteristics.

Incident TB was also more common among PLWH than among people without HIV (IRR 7.37, 95%CI 1.8– 30.7), with incidence rates of 3,538 (95%CI 456–6,621) and 480 (95%CI 0–1,023) per 100,000 person-years, respectively, corresponding to a risk difference of 3,058 (95%CI -71–6188) per 100,000 person-years. Incident TB rates among adult household contacts did not differ by prior TB history (IRR 2.70, 95%CI 0.65–11.25), or by baseline IGRA status, either overall (IRR 0.46, 95%CI 0.11–1.91), or when restricted to the 157 individuals with abnormal CXR (IRR 0.58, 95%CI 0.06–5.57; p=0.64).

Sensitivity analyses that treated participants with negative Xpert Ultra and no MGIT culture result, and participants with trace-positive Xpert Ultra and negative MGIT culture result, as negative, yielded similar results (**Supplementary Figure 1; Supplementary Tables 2 and 3**). A second sensitivity analysis that included participants with clinical (treated) but unconfirmed TB, as incident TB, also showed higher TB incidence rates among individuals with abnormal CXR than those with normal CXR, with incidence rates of 3,931 (95%CI 817– 7,045) and 978 (95%CI 196–1,760) per 100,000 person-years, respectively, corresponding to an IRR of 4.02 (95%CI 1.30–12.39; p=0.015) and a risk difference of 2,953 (95%CI -257–6,164) per 100,000 person-years.

Analysis of incidence rates was further stratified by baseline symptom status. Among the 118 (14.8%) symptomatic participants, abnormal CXR was observed in 25/118 (21.2%) and incident TB developed in 0/25 (0%); normal CXR was observed in 93/118 (78.8%) and incident TB developed in 1/93 (1.1%). TB incidence rates among symptomatic participants with abnormal and normal baseline CXR were zero and 1,140 (95%CI 0–3,381) per 100,000 person-years, respectively; and therefore, IRR could not be computed.

Among the 677 (85.2%) asymptomatic participants, 132/677 (19.5%) had abnormal CXR and incident TB developed in 4/132 (3.03%); normal CXR was observed in 545/677 (80.5%) and incident TB developed in 3/545 (0.55%). TB incidence rates among asymptomatic participants with abnormal and normal baseline CXR were 3,118 (95%CI 93–6,144) and 571 (95%CI 1–1,216) per 100,000 person-years, respectively, corresponding to an IRR of 5.47 (95%CI 1.23–24.25; p=0.025) and a risk difference of 2548 (95%CI -545– 5,641) per 100,000 person-years.

TB incidence rates in symptomatic and asymptomatic participants with normal baseline CXR were 1,140 (95%CI 1–3,374) and 571 (95%CI 1–1,216) per 100,000 person-years, respectively, corresponding to an IRR of 2.00 (95%CI 0.21–19.19; p=0.549) and a risk difference of 569 (95%CI -1756–2,894) per 100,000 person-years.

## Discussion

Our findings add to a growing body of evidence that individuals with known TB exposure and an abnormal CXR suggestive of TB represent a clinically important, high-risk group for progression to TB disease, despite absence of symptoms and negative sputum microbiology at baseline [7, 11, 15]. Over 12 months, close contacts with abnormal CXR had four-fold higher risk of developing microbiologically-confirmed TB than those with normal baseline CXR.

This finding is consistent with the view that some radiographic abnormalities reflect early, paucibacillary disease that is not captured by initial sputum testing. The frequency of baseline CXR abnormalities was similar among asymptomatic and symptomatic participants. However, no incident TB cases were observed in symptomatic participants with abnormal CXR at baseline, which may be due to prior diagnosis of prevalent TB. By contrast, seven (87.5%) of the eight TB progressors were asymptomatic at baseline and, among asymptomatic individuals, those with abnormal CXR had five-fold higher incidence of incident TB, compared to those with normal CXR. This high-risk CXR-positive, sputum-negative phenotype would not usually meet the therapeutic treatment threshold in the absence of TB symptoms, highlighting a potential gap in current management guidelines.

Baseline CXR abnormalities were common in this cohort. Approximately one in five adult household contacts without prevalent TB had an abnormal baseline CXR, which was strongly associated with older age and prior TB, and likely reflects a mix of post-TB radiological sequelae, cumulative environmental and infectious exposures, and age-related structural lung changes, in addition to active disease. This finding complicates interpretation of CXR screening in older adults during contact investigation and underscores the need to distinguish between stable residual changes and features suggestive of active or evolving TB.

This prospective analysis also confirms the major contribution of HIV infection to incident TB risk in adults with close TB exposure. By contrast, baseline IGRA status did not further amplify short-term TB risk among household contacts in this high-incidence setting, even among the 157 individuals with abnormal CXR, although definitive interpretation is constrained by the number of incident TB outcomes. While IGRA positivity is associated with increased TB risk in many studies[16, 17], IGRA has limitations as an individual-level TB predictor in high-transmission settings, where widespread Mtb sensitisation substantially reduces positive predictive value over short time horizons [18].

These results have practical and policy implications for programmatic TB contact investigation in high-burden settings. Symptom and CXR screening are often used to trigger definitive sputum microbiological investigation, but our findings highlight the risk of excluding asymptomatic, CXR-positive, sputum-negative individuals from continued TB disease surveillance. Rather than viewing isolated radiographic findings as false positive, due to other pathology, such individuals should be considered a priority group for enhanced follow-up and more sensitive re-evaluation strategies [19-21]. Options include scheduled symptom review or expanded sputum diagnostic testing, in addition to TPT for known close contacts.

Our findings of increased TB risk in individuals with CXR changes suggestive of TB, despite negative symptom screen and sputum microbiology, are consistent with a meta-analysis of individual participant data from the pre-chemotherapy era [7], a large community TB screening study in China [17], and data from household contacts in Peru [11]. Modelling studies have projected that treating everyone in the community with CXR abnormalities, but without microbiological confirmation of TB, would achieve a sustained decline in TB prevalence to below 50 per 100,000 persons within three annual rounds of population-wide CXR screening in Vietnam [22]. In a large study conducted in Scotland, a single round of mass CXR screening resulted in a major and sustained reduction in incident TB notifications [23]. However, the vast majority (97.5%) of close contacts with abnormal CXR in this study remained well, did not progress to TB over median 12-month follow-up, and would not have benefitted from an additional therapeutic intervention. It follows that indiscriminate treatment of this group risks unnecessary toxicity, avoidable costs, and poor acceptability. Therefore, we agree with the authors of the ICE-TB classification of early TB states that current imaging approaches, in this case CXR, are not specific for TB and might result in overtreatment [24]. The need to balance benefit and risk illustrates the need for more nuanced algorithms that integrate radiological patterns, clinical features (including HIV and prior TB), and local epidemiology to better separate those with evolving TB disease from those with stable residual radiological changes.

Current WHO recommendations state that all household contacts of patients with TB should receive TPT [25]. However, uptake was very low in this cohort. Only 14/795 (1.7%) participants reported receiving TPT, including eight at enrolment and six during follow-up, none of whom progressed to incident PTB. These findings are consistent with delayed implementation of South African TPT guidelines for household contacts of all ages, which were published in February 2023, several months after study enrolment had been completed [26]. The potential benefit of TPT is illustrated by a community-based, randomised controlled trial among IGRA-positive individuals with CXR abnormalities, which demonstrated 55% protective efficacy [27]. By contrast, among a high-risk group categorised by a host blood transcriptomic biomarker, the CORTIS trial did not show efficacy of a three-month TPT regimen for prevention of incident PTB beyond six months [13], casting doubt on whether a preventive, rather than therapeutic treatment regimen is sufficient to halt progression of early, evolving TB disease.

Our study has several limitations. We observed site-level differences in the prevalence of abnormal CXR, which may reflect heterogeneity in local TB burden and Mtb exposure intensity, differing environmental risk profiles, or variability in radiographic reading. Although, the CXR readers were investigators with experience of TB diagnosis, the findings strengthen the case for standardised reading and where feasible, use of computer-aided detection (CAD) to improve consistency across sites. The findings may not generalise beyond South African settings, given the very high IGRA-positive prevalence. In addition, despite the large sample size, the small number of incident TB events limited precision and constrained multivariable modelling of TB progression risk, reflected by wide confidence intervals, especially in sub-group analyses. Consequently, the absolute risk difference of incident TB between abnormal and normal baseline CXR crossed the null threshold and thus was not statistically significant. Therefore, the relative association between abnormal baseline CXR and subsequent TB should be interpreted cautiously. Investigations for incident TB were symptom-triggered, thus asymptomatic incident TB would have been missed and the true burden of incident TB is likely underestimated. Larger longitudinal cohorts, with universal sputum collection for detection of asymptomatic incident TB, would be needed to refine TB risk estimates, explore effect modification (including by HIV and prior TB), and evaluate whether specific radiographic patterns improve prediction. However, our findings persisted in sensitivity analyses that included more participants, with less stringent inclusion criteria for negative sputum microbiology.

In summary, observation of an abnormal baseline CXR suggestive of TB among asymptomatic adult household contacts identified a subgroup at meaningfully increased relative risk of developing microbiologically-confirmed, incident TB over 12 months. These data support incorporation of CXR findings into TB risk stratification frameworks for contact investigation; and warrant prioritisation of this group for careful reassessment, implementation of TPT were indicated, and enhanced surveillance, alongside continued scale-up of antiretroviral therapy and TPT for PLWH. However, most individuals with baseline CXR abnormalities suggestive of TB remained well; and did not develop TB despite very low TPT uptake. It follows that therapeutic TB treatment for this group is not justified, since most recipients would be exposed only to the known potential risks and burdens of treatment, whereas potential benefits would be constrained to a very small number of TB progressors who might be identified by extended surveillance. Future work should focus on better phenotyping of CXR abnormalities and the refinement of guidelines for proportionate, evidence-based management of this group at high risk of TB in high-burden settings.

## Data Availability

The data that support the findings of this study are available on request from the corresponding author. The data are not publicly available because of international data privacy regulations and ethical restrictions.

## Acknowledgements

The authors gratefully acknowledge the study participants and funders.

## Author contributions

TJS, TRS, and MH conceived the idea, raised funds, and provided resources. GW, TJS, TRS, and MH wrote the study protocol and provided study oversight. TM, MT, STM, and FN were responsible for all site-level activities, including recruitment, clinical management, and data collection. KHa provided operational support and project management. HM, FM, RP, KHl, and KS cleaned and verified the underlying data. HM and EM analysed the data. HM, EM, SCM, and MH interpreted the results and wrote the first draft. All authors had full access to the data, confirm the integrity of the data and its presentation, agree with its interpretation as discussed in the manuscript, and reviewed, revised, and approved the manuscript before submission. The corresponding author had final responsibility for the decision to submit for publication.

### Financial support

This research was supported by the RePORT South Africa network with funds received from CRDF Global (University of Cape Town, G-DAA3-19-66875-1; Vanderbilt University Medical Center, G-DAA9-20-66870-1; Stellenbosch University, G-DAA9-20-66918-1; and Wits Health Consortium, G-DAA9-20-66878-1), the NIH (Stellenbosch University, U01AI152075), and the SAMRC. The funders had no role in study design, data collection, analysis, interpretation, the decision to publish, or the preparation of the manuscript.

