## Supplemental Figure S1 and Tables S1, S2 and S3 for "Risk of progression to pulmonary tuberculosis among household contacts with chest radiographic abnormalities in South Africa"

| Name | Affiliation |
| --- | --- |
| Alasdair Leslie | Africa Health Research Institute |
| Farina Karim | Africa Health Research Institute |
| Gareta Dickman | Africa Health Research Institute |
| Gregory Ording-Jespersen | Africa Health Research Institute |
| Kevyna Chetty | Africa Health Research Institute |
| Lindiwe Madziwa | Africa Health Research Institute |
| Thandeka Khoza | Africa Health Research Institute |
| Theresa Smit | Africa Health Research Institute |
| Zainab Baig | Africa Health Research Institute |
| Carolina Mehaffy | Colorado State University |
| John Belisle | Colorado State University |
| Karen Dobos | Colorado State University |
| Kimberly Shelton | Colorado State University |
| ACE Carstens | Department of Biomedical Sciences, Stellenbosch University |
| Andriette Hiemstra | Department of Biomedical Sciences, Stellenbosch University |
| Ané Kruger | Department of Biomedical Sciences, Stellenbosch University |
| Belinda A Kriel | Department of Biomedical Sciences, Stellenbosch University |
| Bronwyn Smith | Department of Biomedical Sciences, Stellenbosch University |
| Candice MacDonald | Department of Biomedical Sciences, Stellenbosch University |
| Candice Snyders | Department of Biomedical Sciences, Stellenbosch University |
| Gian van der Spuy | Department of Biomedical Sciences, Stellenbosch University |
| Ilana R van Rensburg | Department of Biomedical Sciences, Stellenbosch University |
| Ilze Louw | Department of Biomedical Sciences, Stellenbosch University |
| Jane Shaw | Department of Biomedical Sciences, Stellenbosch University |
| Marika Flinn | Department of Biomedical Sciences, Stellenbosch University |
| Susanne Tonsing | Department of Biomedical Sciences, Stellenbosch University |
| Tracy Richardson | Department of Biomedical Sciences, Stellenbosch University |
| Léanie Kleynhans | Department of Biomedical Sciences, Stellenbosch University; Mater Research Institute – The University of Queensland |
| Heather Zar | Department of Paediatrics and Child Health, University of Cape Town |
| Aliasgar Esmail | Division of Pulmonology, Department of Medicine, Groote Schuur Hospital and University of Cape Town Lung Institute |
| Charity Dire | Perinatal HIV Research Unit, University of Witwatersrand |
| Floris Swanepoel | Perinatal HIV Research Unit, University of Witwatersrand |
| Kennedy Otwombe | Perinatal HIV Research Unit, University of Witwatersrand |
| Lorraine Lichakane | Perinatal HIV Research Unit, University of Witwatersrand |
| Lorraine Thobakgale | Perinatal HIV Research Unit, University of Witwatersrand |
| Lyle van de Berg | Perinatal HIV Research Unit, University of Witwatersrand |
| Mbusiseni Ngema | Perinatal HIV Research Unit, University of Witwatersrand |
| Pattamukkil Abraham | Perinatal HIV Research Unit, University of Witwatersrand |
| Thabang Moloja | Perinatal HIV Research Unit, University of Witwatersrand |
| Angelique Luabeya | South African Tuberculosis Vaccine Initiative, University of Cape Town |
| Angelique Mouton | South African Tuberculosis Vaccine Initiative, University of Cape Town |
| Ashley Veldsman | South African Tuberculosis Vaccine Initiative, University of Cape Town |
| Carmen Segelaar | South African Tuberculosis Vaccine Initiative, University of Cape Town |
| Denis Awany | South African Tuberculosis Vaccine Initiative, University of Cape Town |
| Fajwa Opperman | South African Tuberculosis Vaccine Initiative, University of Cape Town |
| Habibullah Valley | South African Tuberculosis Vaccine Initiative, University of Cape Town |
| Hadn Africa | South African Tuberculosis Vaccine Initiative, University of Cape Town |
| Hlengiwe Nkambule | South African Tuberculosis Vaccine Initiative, University of Cape Town |
| Johanna E van Rooyen | South African Tuberculosis Vaccine Initiative, University of Cape Town |
| Justin Shenje | South African Tuberculosis Vaccine Initiative, University of Cape Town |
| Lebohang Makhethe | South African Tuberculosis Vaccine Initiative, University of Cape Town |
| Marcia Steyn | South African Tuberculosis Vaccine Initiative, University of Cape Town |

| | |
|---|---|
| Marwou de Kock | South African Tuberculosis Vaccine Initiative, University of Cape Town |
| Masooda Kaskar | South African Tuberculosis Vaccine Initiative, University of Cape Town |
| Mzwandile Erasmus | South African Tuberculosis Vaccine Initiative, University of Cape Town |
| Nicole Bilek | South African Tuberculosis Vaccine Initiative, University of Cape Town |
| Nicolette Tredoux | South African Tuberculosis Vaccine Initiative, University of Cape Town |
| Nobulumko Khomba | South African Tuberculosis Vaccine Initiative, University of Cape Town |
| Onke Nombida | South African Tuberculosis Vaccine Initiative, University of Cape Town |
| Petrus Tyambetyu | South African Tuberculosis Vaccine Initiative, University of Cape Town |
| Sandisiwe Mangali | South African Tuberculosis Vaccine Initiative, University of Cape Town |
| Sarah Nyangu | South African Tuberculosis Vaccine Initiative, University of Cape Town |
| Simbarashe Mabwe | South African Tuberculosis Vaccine Initiative, University of Cape Town |
| Yolundi Cloete | South African Tuberculosis Vaccine Initiative, University of Cape Town |
| Sara Suliman | University of California San Francisco |
| Lesley Workman | University of Cape Town |
| Linda Mbuthini | University of Cape Town |
| Lindsay Wilson | University of Cape Town |
| Ryan Johnson | University of Cape Town |
| Tahira Kootbodien | University of Cape Town |
| Andrea Kotze | University of Cape Town Lung Institute |
| Cynthia Baard | University of Cape Town Lung Institute |
| Keertan Dheda | University of Cape Town Lung Institute |
| Rodney Dawson | University of Cape Town Lung Institute |
| Shameem Jaumdally | University of Cape Town Lung Institute |
| Tahlia Perumal | University of Cape Town Lung Institute |
| Bernard Fourie | University of Pretoria |
| Gerard Cangelosi | University of Washington |
| Rachel C Wood | University of Washington |
| Hilary Vansell Riley | Vanderbilt University Medical Center |
| Marina Cruvinel Figueiredo | Vanderbilt University Medical Center |
| Megan Turner | Vanderbilt University Medical Center |
| Rebecca Berhanu | Vanderbilt University Medical Center |
| Stephany Norah Duda | Vanderbilt University Medical Center |
| Travis Harris | Vanderbilt University Medical Center |

### Supplementary tables

**Table S1:** Characteristics of participants who progressed to incident pulmonary TB.

| Variable | Total | Abormal CXR | Normal CXR |
|---|---|---|---|
| | N=8 | N=4 | N=4 |
| Age, median (IQR) | 39.2 (35.1–51.9) | 51.9 (42.6–56.7) | 36.9 (33.5–39.2) |
| Female sex, n (%) | 7 (87.5) | 4 (100) | 3 (75) |
| Ancestry, n (%) | | | |
| Black African | 4 (50) | 1 (25) | 3 (75) |
| Mixed | 4 (50) | 3 (75) | 1 (25) |
| Caucasian | 0 (0) | 0 (0) | 0 (0) |
| Smoking history, n (%) | | | |
| Never | 4 (50) | 2 (50) | 2 (50) |
| Current smoker | 4 (50) | 2 (50) | 2 (50) |
| Former smoker | 0 (0) | 0 (0) | 0 (0) |
| Prior TB, n (%) | 3 (37.5) | 3 (75) | 0 (0) |
| HIV positive, n (%) | 5 (62.5) | 2 (50) | 3 (75) |
| IGRA, n (%) | | | |
| Negative | 3 (37.5) | 1 (25) | 2 (50) |
| Positive | 5 (62.5) | 3 (75) | 2 (50) |
| Study Site, n (%) | | | |
| Klerksdorp | 3 (37.5) | 1 (25) | 2 (50) |
| Worcester | 5 (62.5) | 3 (75) | 2 (50) |
| Ravensmead | 0 (0) | 0 (0) | 0 (0) |
| TB Symptoms, n (%) | 1 (12.5) | 0 (0) | 1 (25) |
| TB preventive therapy, n (%) | 0 (0) | 0 (0) | 0 (0) |
| Person-time (months) | 8.2 (6.7–12.1) | 10.1 (7–13.5) | 8 (6.4–10.2) |

The table presents baseline characteristics of the 8 individuals who progressed to incident TB (tuberculosis). IQR, interquartile range. IGRA, interferon-γ release assay. CXR, chest radiograph.

**Table S2:** Baseline characteristics and factors associated with an abnormal baseline chest radiograph (sensitivity analysis).

| Variable | Total | Abnormal CXR | Normal CXR | Univariable Analysis | | Multivariable Analysis | |
|---|---|---|---|---|---|---|---|
| | N=850 | N=173 | N=677 | OR (95% CI) | P-Value | aOR (95% CI) | P-Value |
| Age, median (IQR) | 35.0 (25.6–48.2) | 43.0 (30.5–53.5) | 33.7 (24.8–45.8) | 1.03 (1.02–1.04) | <0.001 | 1.04 (1.02–1.05) | <0.001 |
| Female sex, n (%) | 547 (64.4) | 103 (59.5) | 444 (65.6) | 0.77 (0.55–1.09) | 0.14 | 0.74 (0.49–1.12) | 0.16 |
| Ancestry, n (%) | | | | | | | |
| Black African | 401 (47.2) | 53 (30.6) | 348 (51.4) | Reference | – | Reference | – |
| Mixed | 446 (52.5) | 119 (68.8) | 327 (48.3) | 2.39 (1.67–3.42) | <0.001 | 1.24 (0.73–2.12) | 0.42 |
| Caucasian | 3 (0.4) | 1 (0.6) | 2 (0.3) | 3.28 (0.29–36.89) | 0.34 | 2.18 (0.09–50.80) | 0.63 |
| Smoking history, n (%) | | | | | | | |
| Never | 356 (41.9) | 52 (30.1) | 304 (44.9) | Reference | – | Reference | – |
| Current smoker | 449 (52.8) | 109 (63.0) | 340 (50.2) | 1.87 (1.30–2.70) | 0.001 | 1.56 (0.99–2.48) | 0.057 |
| Former smoker | 45 (5.3) | 12 (6.9) | 33 (4.9) | 2.13 (1.03–4.38) | 0.041 | 1.90 (0.95–3.79) | 0.068 |
| Prior TB, n (%) | 158 (18.6) | 79 (45.7) | 79 (11.7) | 6.36 (4.35–9.31) | <0.001 | 6.44 (4.31–9.62) | <0.001 |
| HIV positive, n (%) | 159 (18.7) | 33 (19.1) | 126 (18.6) | 1.03 (0.67–1.57) | 0.91 | – | – |
| IGRA, n (%) | | | | | | | |
| Negative | 171 (20.1) | 27 (15.6) | 144 (21.3) | Reference | – | – | – |
| Positive | 620 (72.9) | 130 (75.1) | 490 (72.4) | 1.41 (0.90–2.23) | 0.13 | – | – |
| Indeterminate | 33 (3.9) | 9 (5.2) | 24 (3.5) | 2.00 (0.84–4.77) | 0.12 | – | – |
| Not done | 26 (3.1) | 7 (4.0) | 19 (2.8) | 1.96 (0.75–5.13) | 0.17 | – | – |
| Study site, n (%) | | | | | | – | – |
| Klerksdorp | 309 (36.4) | 33 (19.1) | 276 (40.8) | Reference | – | Reference | – |
| Worcester | 386 (45.4) | 88 (50.9) | 298 (44.0) | 2.47 (1.60–3.81) | <0.001 | 2.05 (1.09–3.84) | 0.026 |
| Ravensmead | 155 (18.2) | 52 (30.1) | 103 (15.2) | 4.22 (2.58–6.90) | <0.001 | 5.02 (2.49–10.10) | <0.001 |
| TB symptoms, n (%) | 126 (14.8) | 27 (15.6) | 99 (14.6) | 1.08 (0.68–1.72) | 0.75 | – | – |
| TB preventive therapy at baseline, n (%) | 8 (0.9) | 0 (0) | 8 (1.2) | – | – | – | – |

The table presents baseline characteristics and factors associated with an abnormal baseline chest radiograph in the sensitivity analysis. The sensitivity analysis included participants who were excluded from the primary analysis because of uncertain baseline TB status: those with a negative Xpert Ultra result but missing or contaminated MGIT culture, and those with trace-positive Xpert Ultra but negative MGIT culture. For this sensitivity analysis, these participants were classified as TB negative at baseline. All p values were computed using logistic regression. IQR, interquartile range. OR, odds ratio. aOR, adjusted odds ratio. CI, confidence interval. CXR, chest radiograph. TB, tuberculosis. IGRA, interferon-γ release assay.

**Table S3:** Risk of progression to incident pulmonary TB by baseline characteristics (sensitivity analysis).

| Variable | Confirmed TB / N participants N=850 | Incidence rate per 100,000 person-years (95% CI) | Incidence rate ratio (95% CI) | Risk difference per 100,000 person-years (95% CI) |
|---|---|---|---|---|
| Chest radiography | | | | |
| Normal | 4/677 | 617 (13–1221) | Reference | Reference |
| Abnormal | 4/173 | 2399 (66–4731) | 3.89 (1.3–11.99) | 1782 (-628–4191) |
| Sex | | | | |
| Male | 1/303 | 343 (-329–1014) | Reference | Reference |
| Female | 7/547 | 1338 (350–2325) | 3.91 (0.48–31.73) | 995 (-199–2189) |
| Ancestry | | | | |
| Black African | 4/401 | 1066 (23–2109) | Reference | Reference |
| Mixed | 4/446 | 915 (20–1810) | 0.86 (0.22–3.42) | -151 (-1525–1223) |
| Caucasian | 0/3 | 0 | – | – |
| Smoking history | | | | |
| Never | 4/356 | 1188 (26–2349) | Reference | Reference |
| Current smoker | 4/449 | 918 (20–1815) | 0.77 (0.19–3.08) | -270 (-1738–1198) |
| Former smoker | 0/45 | 0 | – | – |
| Prior TB | | | | |
| No | 5/692 | 754 (94–1413) | Reference | Reference |
| Yes | 3/158 | 1977 (-250–4205) | 2.62 (0.63–10.93) | 1224 (-1099–3547) |
| HIV status | | | | |
| Negative | 3/691 | 451 (-59–960) | Reference | Reference |
| Positive | 5/159 | 3344 (430–6259) | 7.42 (1.78–30.92) | 2894 (-65–5852) |
| IGRA | | | | |
| Negative | 3/171 | 1843 (-238–3923) | Reference | Reference |
| Positive | 5/620 | 841 (106–1577) | 0.46 (0.11–1.9) | -1002 (-3208–1206) |
| Indeterminate | 0/33 | 0 | – | – |
| Not done | 0/26 | 0 | – | – |
| Study Site | | | | |
| Klerksdorp | 3/309 | 1060 (-137–2257) | Reference | Reference |
| Worcester | 5/386 | 1312 (166–2458) | 1.24 (0.3–5.16) | 252 (-1405–1909) |
| Ravensmead | 0/155 | 0 | – | -1060 (-2257–137) |
| Symptomatic | | | | |
| No | 7/724 | 1006 (263–1749) | Reference | Reference |
| Yes | 1/126 | 837 (-803–2478) | 0.83 (0.1–6.76) | -169 (-1970–1632) |
| TB preventive therapy | | | | |
| No | 8/842 | 991 (306–1676) | – | Reference |
| Yes | 0/8 | 0 | – | -991 (-1676—306) |

The table presents TB incidence rates, relative risks (incidence rate ratios), and absolute risk differences (attributable risk) by baseline characteristics in the sensitivity analysis. The sensitivity analysis included participants who were excluded from the primary analysis because of uncertain baseline TB status: those with a negative Xpert Ultra result but missing or contaminated MGIT culture, and those with trace-positive Xpert Ultra but negative MGIT culture. For this sensitivity analysis, these participants were classified as TB negative at baseline. Incidence rate ratios were computed using Poisson regression. The absolute risk difference is computed as the difference in risk between the presence and absence of a specific exposure variable. IR, incidence rate. IRR, incidence rate ratio. CI, confidence interval. TB, tuberculosis. IGRA, interferon-γ release assay.

### Supplementary figures

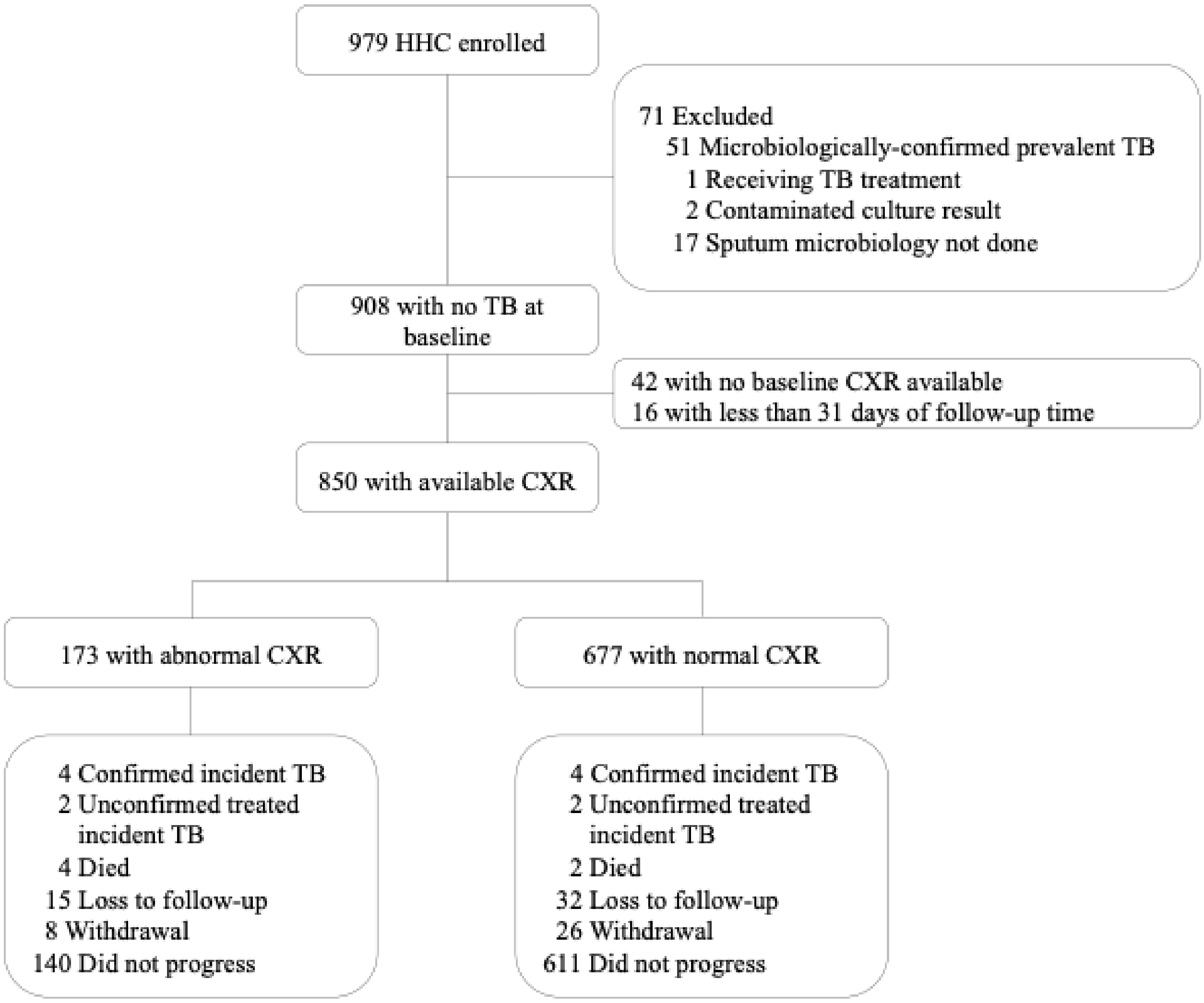

**Figure S1:** Study flow for the participants included in the sensitivity analysis.

Study flow diagram detailing exclusions, chest radiograph results, and follow-up outcomes for the sensitivity analysis. Unconfirmed, treated incident TB denotes participants who were clinically diagnosed with TB without microbiological confirmation and started therapeutic TB treatment. HHC, household contact. TB, tuberculosis. CXR, chest radiograph.
